# *I am a quarterback:* A mixed methods study of death investigators’ communication with family members of young sudden cardiac death victims from suspected heritable causes

**DOI:** 10.1101/2024.09.13.24313665

**Authors:** Katherine L. Mason, Katherine S. Allan, Dirk Huyer, June Carroll, Arnon Adler, Julie Rutberg, Sheldon Cheskes, Steve Lin, Erik Mont, Lindsay Denis, Joel A. Kirsh, Kris Cunningham, Jodi Garner, Liz Siydock, Katie N. Dainty, Matthew Bowes, Karolyn Yee, Paul Dorian, Krystina B. Lewis

**Affiliations:** School of Nursing, Faculty of Health Sciences, University of Ottawa, Ottawa, Ontario, Canada; Division of Cardiology, Unity Health Toronto – St. Michael’s Hospital, Toronto, Ontario, Canada; Office of the Chief Coroner of Ontario, Toronto, Ontario, Canada; Department of Medicine, University of Toronto, Toronto, Ontario, Canada; Department of Pediatrics, University of Toronto, Toronto, Ontario, Canada; Department of Family & Community Medicine, Sinai Health, University of Toronto, Toronto, Ontario, Canada; Department of Cardiology, Toronto General Hospital, Toronto, Ontario, Canada; Children’s Hospital of Eastern Ontario (CHEO), Ottawa, Ontario, Canada; Nova Scotia Medical Examiner Service, Dartmouth, Nova Scotia, Canada; Ontario Forensic Pathology Service, Toronto, Ontario, Canada; Family Member, Toronto, Ontario, Canada; Patient Centred Outcomes, North York General Hospital, Toronto, Ontario, Canada; University of Ottawa Heart Institute, Ottawa, Ontario, Canada

**Keywords:** Sudden cardiac death, heritable cardiac condition, cause of death, communication, coroner, death investigator, mixed methods

## Abstract

**Background:** Sudden cardiac death (SCD) is a devastating event and a leading cause of mortality, globally. In the young (2-45 years), SCD is often attributable to a heritable cardiac condition. Death investigators are often responsible for investigating the cause of death and communicating their results and risk of heritable cardiac conditions with family members of SCD victims. Family often struggles to comprehend the information that is communicated to them.

**Purpose:** To understand the delivery, reach and impact of communication strategies informing family members of SCD victims about their relative’s cause of death and their own risk for heritable cardiac conditions.

**Methods:** We conducted an explanatory sequential mixed methods study. We collected quantitative data via a web-based survey and qualitative data via telephone interviews to investigate how death investigators in Ontario and Nova Scotia, Canada, communicate with family members of SCD victims. We used descriptive statistics to analyze the survey data and thematic analysis to analyze the qualitative data. We triangulated data at multiple levels.

**Results:** Between October 2022 and July 2023, we surveyed 78 death investigators and interviewed a subset (n=20). Death investigators reported that SCDs due to suspected heritable cardiac conditions were more difficult (40%, n=31) or slightly more difficult (35%, n = 27) to investigate, often requiring a higher frequency of communication with families. Death investigators reported contacting family members via phone (n=75, 96.1%) and used various strategies to achieve their communication goals. Strategies were influenced by family characteristics; involvement of other professionals; characteristics of the investigation, access to resources, and system-level barriers.

**Conclusion:** SCD investigations in the young due to suspected heritable cardiac conditions were more challenging and required a higher frequency of communication. Death investigators used various strategies to achieve their communication goals. Further research should examine how systematic changes can improve communication with family members.

**What is Known?:**

- Sudden cardiac death (SCD) is a devastating and unexpected event that can be caused by heritable cardiac conditions, putting the decedents family at risk of SCD.
- Communication with death investigators and other health care professionals influences families’ experiences learning about the cause of death and about their risk for heritable cardiac condition.

**What this Study Adds:**

- According to death investigators, SCD cases due to suspected heritable cardiac conditions are more difficult to investigate, require a higher frequency of communication with family members than other types of cases, and benefit from using different communication modalities.
- Despite their best intentions, death investigators are contending with many factors beyond their control that influence how communication with family members is carried out.
- Provincial death investigation systems alone do not currently provide families of SCD victims with sufficient communication, as families often seek external resources.

## Background

Sudden cardiac death (SCD) is a devastating event that remains a leading cause of mortality globally.^1^ SCD is defined as a natural event, characterized by cardiorespiratory collapse that occurs suddenly, unexpectedly, and is presumed to be due to a cardiac cause.^1,2^ In many healthcare systems around the world, legislation mandates the investigation of sudden and unexpected deaths.^3,4^ Death investigators, including coroners, medical examiners and coordinators of investigations (henceforth referred to as *death investigators*), are responsible for communicating with bereaved family members about the cause of death and their potential risk for heritable cardiac conditions, particularly in cases where the victim is young (2-45 years old).

In the early days and months following the SCD, family members are informed of the death investigation process, the cause of death, and their own risk for heritable cardiac conditions. The way in which this is communicated is critical, as it shapes the families’ experiences during this difficult time and may influence their choice of pursuing subsequent cardiac screening and genetic testing, when recommended.^5^ Policies, procedures, and practices for death investigations vary by jurisdiction.^3^ Communication-specific training is often dependant on the individual death investigator’s professional experiences in their other specialties and their own motivations, creating variability and inconsistency in approaches within and across death investigation systems.^5^ In addition, many families describe their struggles with understanding and remembering information during the emotional and overwhelming period following the sudden death of a loved one.^5^

Despite the importance for bereaved family members to receive effective communication, there is little research investigating what constitutes optimal communication in this context. In fact, little is known about the communication approaches in place within death investigation systems. This study seeks to explore how death investigators communicate with families of young SCD victims about the cause of death and their risk for SCD, and what influences their communication approaches. Three research questions guided this study:

1. What is the type and timing of communication strategies used by death investigators when communicating cause of death and the risk of SCD with families of young SCD victims from suspected heritable causes (Quantitative)?
2. What are the experiences of death investigators’ communication about the cause of death and risk of SCD with families of young SCD victims from suspected heritable causes, and what recommendations do they suggest to improve communication (Qualitative)?
3. How does the integration of qualitative and quantitative findings enhance our understanding of death investigators’ experiences with communication during investigations of young SCD victims from suspected heritable causes (Quantitative and Qualitative)?

## Methods

### Study Design

We conducted a sequential mixed methods explanatory sequential study to explore how death investigators communicate with families of young SCD victims about the cause of death and their risk for SCD.^6^ We collected quantitative data via a web survey, which informed the interview guide. The interview guide was used to collect qualitative data via individual interviews. The study was approved by the University of Ottawa Research Ethics Board (H-09-21-7135). We report this study using the CHERRIES^7^ and SRQR^8^ reporting guidelines.

### Research Characteristics and Reflexivity

Members of the Office of the Chief Coroner of Ontario (D.H., K.C., J.A.K., L.S.) and Nova Scotia Medical Examiner Service (M.B., E.M., L.D.) including death investigators and the Family Genetic Care Associate (K.S.A.; co-first author) were interested in learning about current communication approaches and the factors influencing them to inform a future evidence-informed approach to improve communication, supports, and resources offered to families in practice. A family member partner (J.G.) with lived experience of losing a young family member to SCD was motivated to collaborate to improve the lengthy death investigation process for anyone having to navigate these tremendously challenging waters. Our research team was comprised of early to senior career investigators (K.B.L., K.S.A., K.N.D.), including trainees (K.L.M., K.Y.), clinician-researchers in Cardiology, (A.A., P.D.), Emergency Medicine (S.L.) Family Medicine (J.C., S.C.), genetic counsellor (J.R.), all with expertise, interest, and recognition of the need to improve family-centered communication for informed, values-based care in suspected heritable sudden cardiac death cases. Many members of the research team held content and methodological expertise, yet were outsiders to the death investigation process and could therefore provide an independent and impartial examination of current communication processes and impact of these on death investigators’ experiences. Together, our team committed to an integrated knowledge translation (IKT) approach to the study, an approach that has been widely promoted and used in health research in Canada.^9^ Kothari et al. define IKT as “a model of collaborative research, where researchers work with knowledge users who identify a problem and have the authority to implement the research recommendations”.^10^ Hence, this research was shaped and conducted through collaborations between the research team and knowledge users, from the conceptualization of the research to the dissemination of the findings for greatest impact.

### Setting

This study was conducted in two Canadian provinces, Ontario and Nova Scotia. These sites were selected based on their membership in the Canadian Sudden Cardiac Arrest Network (C-SCAN)^11^, number of autopsies performed yearly in young SCD victims, and their capacity to support the research project. In these provinces, the Office of the Chief Coroner of Ontario, and the Nova Scotia Medical Examiner Service, respectively, are responsible for investigating all sudden and unexpected deaths, supporting families through the death investigation process, and informing them of the results. In Ontario, coroners conduct death investigations in collaboration with forensic pathologists, while in Nova Scotia, medical examiners and coordinators of investigation work collaboratively. In this paper, we use the term death investigators to refer to coroners, medical examiners, and coordinators of investigations.

### Eligibility Criteria

We invited death investigators working in Ontario and Nova Scotia, who had investigated at least one young (ages 2-45) SCD case between 2018 and 2021, where the death was attributable to a heritable cardiac condition, or a case with no anatomical or toxicological cause that was presumably due to a heritable cardiac condition. Heritable cardiac conditions were defined according to the Canadian Cardiovascular Society Guidelines.^12^ Participants were required to read, speak, and understand either English or French.

### Recruitment

All Ontario (n=340) and Nova Scotia (n=19) death investigators were invited to participate in the anonymous web survey by email through their institutional staff list servs. Two email reminders were sent to all eligible participants approximately one month apart, in keeping with Dillman’s survey methods.^13^ Once the survey was completed, respondents could indicate their interest in participating in a subsequent telephone interview by providing their name and email. We contacted interested interview participants by email with one follow-up reminder, if required.

### Data collection

#### Quantitative data collection

We collected quantitative data via an anonymous web-survey, using Hosted in Canada Surveys between October 2022 and February 2023. The secure link was included in the study invitation email. Participants were informed of the length of the survey, where the data was stored, the investigators and the purpose of the study on the first page of the web survey. Survey completion implied informed consent. The research team developed the survey based on known gaps revealed from our prior work aligned with the study aims,^2^ and the literature on communication.^5^ The web survey was piloted by members of the research team, including a Regional Supervising Coroner (J.K.) and revised for clarity, flow and technical functionality prior to its launch. The web survey was comprised of five sections with both open and closed-ended questions on: 1) sociodemographic characteristics, 2) communication with SCD families (e.g., type, frequency, modality), 3) recommendations for cardiac and/or genetic testing follow-up, 4) interactions with genetic counsellors, and 5) needs for investigating SCD cases. The web survey questions were non-randomized and used adaptive questioning. It was comprised of 37 questions, distributed page-by-page. Participants were able to navigate backwards through the survey to review their answers. During quantitative data collection, data stored securely within the Hosted in Canada database, which adheres to the Personal Information Protection and Electronic Documents Act. Since the completion of the data collection, data is stored in the University of Ottawa Microsoft OneDrive. Web survey participants were provided with a $20 electronic coffee gift card following web survey completion, if they chose to provide an email address. Email addresses were checked for duplicates to prevent multiple entries from the same individual.

#### Qualitative data collection

We used a qualitative descriptive approach. The research team developed a semi-structured interview guide informed by the literature, our team’s prior work in this area^5^, and refined it based on the survey results. Questions were designed to (1) explore death investigators’ experiences of communicating the cause of death and risk of heritable cardiac condition to families, (2) understand the type and timing of communication strategies used with families, and (3) to recognize the role of other professionals in the death investigation process. From March to July 2023, K.S.A, K.B.L and K.L.M conducted telephone interviews with all death investigators who agreed to participate. We audio recorded, transcribed, and de-identified the interviews prior to analysis. Qualitative transcripts are stored in the University of Ottawa Microsoft OneDrive. Participants were provided with an additional $20 electronic coffee gift card following the completion of the interview.

### Data Analysis

#### Quantitative data analysis

Only participants who had completed at least one of the five survey sections were included in our analysis. We used descriptive statistics to analyze the quantitative data. Continuous data were reported as means and standard deviations, while categorical data were reported as counts and percentages.

#### Qualitative data analysis

We used an iterative team-based process led by two PhD-prepared university-based researchers (K.S.A., K.B.L.) and two trainees (K.Y., K.L.M) to analyze all interview transcripts guided by Braun and Clark’s thematic analysis approach.^14,15^ We identified, analyzed, organized, and described themes that reflected death investigators’ experiences of conducting SCD investigations and communicating with families of SCD victims. Each transcript was analyzed independently by at least two team members and discussed as a team on a weekly basis. Meaningful segments of data were coded and organized in a Microsoft Excel-based codebook. As analysis progressed, we revised the codebook and clustered codes into overarching themes. Perspectives were discussed and disagreements in coding were resolved at weekly team meetings. Early transcripts were re-analyzed following the finalization of the codebook to ensure all relevant data was captured. Open-ended survey responses were read to determine if any new themes that were not captured in the codebook emerged.

#### Data integration

We integrated qualitative and quantitative data at multiple levels. At the level of study design, we integrated data through our sequential approach. At the level of the sampling frame, we integrated data by recruiting survey participants for telephone interviews, and we integrated data from our data collection tools by modifying our interview guide based on survey results. We also integrated data in our interpretations of the findings.

## Results

### Participant characteristics

Of 359 invited Ontario and Nova Scotia death investigators 76 participants fully completed and 2 participants partially completely the web-survey (response rate = 78/359, 22%). One hundred and five participants navigated past the first page of the survey (completion rate = 76/105, 72%). Participants were 55 +/− 14 years old on average, 31 (42%) female, 31 (42%) identified as women. Twenty (25%) survey participants, 18 from Ontario and 2 from Nova Scotia, participated in telephone interviews. Interview participants’ characteristics were similar to survey participants, although with less ethnic diversity. Participants were 55 +/− 13 years old on average, with the same proportions of females and women 40% (n = 8). Participants’ characteristics for both the survey and the interviews are detailed in Table I.

**Table I.** Characteristics of death investigators.

|  | **Web Survey Participants**<br>**(n = 78)** | **Interview Participants**<br>**(n = 20)** |
| --- | --- | --- |
| **Age (years)** | 54.9 +/- 13.8 | 54.7 +/- 12.5 |
| **Sex (female)** | 31 (41.9%) | 8 (40.0%) |
| **Gender (women)** | 31 (41.9%) | 8 (40.0%) |
| **Ethnicity** |  |  |
| White | 56 (71.9%) | 18 (90.0%) |
| East Asian | 7 (9.0%) | 2 (10.0%) |
| South Asian | 4 (5.1%) | 0 (0.0%) |
| Middle Eastern | 3 (2.6%) | 0 (0.0%) |
| **Experience as a death investigator** |  |  |
| < 1 year | 3 (3.8%) | 1 (5.0%) |
| 1-4 years | 19 (24.5%) | 5 (25.0%) |
| 5-10 years | 15 (19.2%) | 3 (15.0%) |
| > 10 years | 37 (47.4%) | 11 (55.0%) |
| No answer | 4 (5.1%) | 0 (0.0%) |
| **Number of SCD cases investigated** |  |  |
| 1-5 | 15 (19.2%) | 6 (30.0%) |
| 6-10 | 12 (15.4%) | 6 (30.0%) |
| 11-15 | 6 (7.7%) | 1 (5.0%) |
| > 15 | 26 (33.3%) | 7 (35.0%) |
| No answer | 19 (24.3%) | - |

### Quantitative Web Survey Results

Most death investigators reported SCD cases due to suspected heritable cardiac conditions are more (n=31, 40%) or slightly more (n=27, 35%) difficult to investigate than other cases. They also reported that they require more (n=31, 40%) or slightly more (n=31, 40%) frequency of communication with family members than other types of cases (Tables II and III). To communicate with families, death investigators described using different modalities, either alone or in combination. Their choice of modality depended on: 1) their personal preferences, 2) the circumstances of the investigation, and/or 3) the preferences of the family. Death investigators most often used the phone (n=75, 96%), followed by in-person (n=35, 45%), email (n=32, 41%), with text messages (n=8, 10%) and written letters used less often (n=8, 10%).

**Table II.** Communication with family members of SCD victims.

|  | **n (%)** | **Supporting Quotations** |
| --- | --- | --- |
| **Difficulty of SCD cases compared to other cases.** |  |  |
| More difficult than other cases. | 31 (39.7%) | *"They are more emotionally charged by families as they are less expected."* |
| Slightly more difficult than other cases. | 27 (34.6%) | *"[The investigation is] often less straightforward and [there are] often more questions to address"* |
| About the same as any case. | 18 (23.1%) | *"Every coroner case is a puzzle that needs to be solved, and every family wants answers."* |
| Slightly less difficult than other cases. | 1 (1.3%) | *"Traumatic deaths, suicides and unexplained death circumstances are much more difficult."* |
| A lot less difficult than other cases. | - | None provided. |
| No answer | 1 (1.3%) |  |

**Table III.** Frequency of communication with family members of SCD victims.

|  |  |  |
| --- | --- | --- |
| More frequency of communication than other cases | 31 (39.7%) | *"[There are] more questions to answer and often more people to call."* |
| Slightly more frequency of communication than other cases | 31 (39.7%) | *"At least one or more extra conversations are needed after the genetics come back in order to disclose [risk] and ensure [the family] understands the implications."* |
| About the same as any case | 15 (19.2%) | *"Spending time on the phone with families is more so based on their unique requirements."* |
| Slightly less frequency of communication than other cases | 0 (0.0%) | None provided. |
| A lot less frequency of communication than other cases | 0 (0.0%) |  |
| No answer | 1 (1.3%) |  |

During their investigations, death investigators reported communicating with a variety of experts, including their supervising death investigator, the Family Genetic Care Associate and/or Family Liaison support, family physicians and first responders. They also frequently consulted genetic specialists (n=51, 68%), forensic pathologists (n=44, 59%), cardiac arrhythmia specialists (n=44, 59%) and more experienced death investigators (n=13, 17%). Other experts’ involvement depended on the death investigator’s perceived scope of their own role, their prior experience, the family’s needs, and the characteristics of the case.

Nearly half of surveyed death investigators were aware of recommendations (n=35, 47%) from their institution for investigating SCD cases. Most death investigators who were aware of them described them as very or extremely helpful (n=24, 65%).

### Qualitative Telephone Interviews

In our thematic analysis of interview transcripts, we revealed that participants were intrinsically driven by three primary communication goals, which ultimately drove the selection of the strategies used to achieve them (Theme 1). Yet, a variety of factors influenced their ability to communicate as they’d hope, limiting their ability to communicate effectively (Theme 2), including characteristics of the family, involvement of other professionals, characteristics of the investigation, access to resources, and system-level barriers all of which are presented as sub-themes.

### Theme 1. What are death investigators aiming to communicate – and how

Death investigators reported that their primary goals were (1) to inform the family about the cause of death; (2) to protect the living by adequately informing family members about their risk for heritable cardiac conditions; and (3) to recommend the referral of family members for cardiac follow-up and screening.

Death investigators described numerous verbal and non-verbal strategies to achieve their communication goals, most often drawing on their training and experiences as healthcare professionals to establish rapport with families that works best for them. First, they aimed to communicate with empathy and compassion, expressing their willingness to be open and available to answer family members’ questions, encouraging families to reach out if they needed additional information, and offering reassurance. A death investigator described their desire *“To listen. Show compassion and empathy. To tell them that I am there for them.”* Another expressed that *“it’s just overwhelming grief [for the families] and sometimes they have guilt and so I need to reassure them there isn’t anything they could have done differently.”*

Death investigators discussed the importance of setting expectations for the death investigation before starting the investigation with frequent updates as it is ongoing, which help families prepare for the lengthy timelines and complex logistics. One death investigator described calling the primary contact during an investigation to inform them that “*we haven’t found a cause of death, it’s going to take longer, we’re going to be doing genetic testing and other toxicology. So, I’ll call you again, but please be prepared, it’s going to be maybe three months, maybe longer before you hear from me again.”* Many participants referred to following up with families, repeating information, using plain and clear language and reading reports with family members to enhance family members’ recall and understanding. One death investigator described that communication is often over the phone and *“that’s where the follow-up phone call comes in.” Quite often [families] have a few more questions and I have some more questions too, so it’s a much longer conversation usually”*.

### Theme 2. What influences death investigators’ communication

Death investigators reported numerous factors impacted their selection and perceived usefulness of communication strategies, which prevented some death investigators from communicating in their desired manner. Influencing factors included (1) characteristics of the family; (2) involvement of other professionals; (3) characteristics of the investigation, (4) access to resources, and (5) system-level barriers.

#### (1) Characteristics of the family

Several family-related characteristics influenced the communication strategies that death investigators used. These included differences in the ability of family members to retain and understand information, primarily because of their grief. One death investigator noted that *“we know that people only retain around 10% of what they’re told”* during the period immediately following the sudden death. Some death investigators reported tailoring their language according to families’ level of education and their understanding of medical terminology. One death investigator described the challenge of *“explain[ing] some of the details to the family. And to be technical enough without overwhelming families.”* Some death investigators had the perception that families’ prioritization for the death investigation or seeking screening for themselves varied, as one described that *“people have a lot going on in their lives. Is it because they’re too busy trying to buy groceries and keep a roof over their head and keep their kids in school and this is just number 20 on their list of priorities?”* Additional family-related characteristics were attributable to emotionally heightened circumstances following the death of a loved one and complicated family dynamics with one death investigator who explained that *“there’s a lot of layers to these kinds of conversations. And they can be time consuming.”*

#### (2) Involvement of other professionals

Death investigators spoke about their responsibility managing communication between family members and other experts throughout the investigation. For example, one death investigator described themselves as a “*quarterback”,* who explained they were: “*there to make sure that the various members of the team talk to each other, do their appropriate tasks… I’m not the genetic counselor, the genetic counseling is not up to me. It’s up to someone who’s expert at it. The pathologist provides information. I put it into context. It’s my role to really help coordinate things to ensure families are aware of the resources and to ensure that nothing falls between the cracks.”* When cases did involve multiple other professionals, questions arose about “*who has ownership and responsibility”* of communicating results to families, describing the importance “*of what’s considered the scope of [their] work and what isn’t, because of so many people not having access to family doctors”*.

For others, it was challenging to even know who to engage in each case. For example, one death investigator described their hesitation, saying *“Well, who needs genetic testing and who should they see for that? Do they see a cardiologist? Because I don’t know that the cardiologist in my town would necessarily themselves be able to refer them. I can’t refer them to a geneticist, they need to see their family doctor and have their family doctor refer them to a geneticist. And then I get a call from the family doctor and they’re like, how do I do this? Because they’ve never encountered it before either.”* Some death investigators in Ontario reported directing family members of SCD victims to the Family Genetic Care Associate to facilitate cascade screening, cardiac care, and genetic testing. Others reported directing family members to their family physician, who may assume responsibility for organizing further testing of surviving family members, connect them with specialists, and discuss the death investigation results.

#### (3) Characteristics of the Investigation

Death investigators explained the heightened challenge with suspected heritable cardiac conditions SCD cases as compared to other cases, because the cause of death can be difficult or impossible to determine. This results in an added responsibility for death investigators, who may need to help families navigate unknown circumstances. For example, one death investigator described how *“the idea that even at the end of all the testing there’s sometimes not a satisfying answer that [they] can give to the families. Their disappointment is probably the hardest thing to deal with in these cases”.* The lack of answers can lengthen the process, sometimes *“be upward of a year”* in length, and ultimately leaving both families and death investigators without a known cause of death. For example, one death investigator described this process, saying *“In terms of getting an answer, that requires more phone calls to say, we still don’t really know. We’re going to do these extra tests and that’s going to take months. And that’s really, really hard on these families”*.

#### (4) Access to Resources

Death investigators described how their access to resources, prior medical background, and ongoing professional and skills development shaped their communication with families of SCD victims. For example, one death investigator said: “*I’m a Family Medicine doctor by training, so we do a lot of interviewing and finding out the expectations of families, I think that kind of training of how to communicate has been probably what I draw on the most when I’m talking to the families”*.

Others described having to independently seek their own resources, and professional development opportunities to obtain additional information on certain topics to ensure they were best prepared to provide families with detailed and accurate information. One death investigator described that *“if [they are] going to be providing some of that information to the family, then [they] think [they] need to be clear”*.

Many death investigators were aware of recommendations from their institution for investigating SCD cases: “*a standardized approach, based on best practices, is always a good idea”.* Recommendations were delivered through investigating manuals, courses, and consultations with other experts. Many death investigators expressed the value of additional training on specific types of cardiac conditions, counselling families and the intricacies of referral processes. This includes understanding when and how to contact other experts, including geneticists, genetic counsellors, and medical specialists. Death investigators also suggested new resources that could be useful, including educational documents on SCD, training to use the institutional database, and lists of experts to contact if needed.

#### (5) System Level Barriers

Some participants noted several system-level barriers that hindered the frequency and timing of useful and ethical communication. Identified barriers included gaps of several months from initial contact with families to having results from post-mortem reports. This was deemed a critical barrier, as such delays impacted family member’s referrals to medical specialists for cascade screening to determine their potential risk for SCD due to a heritable cardiac condition. One death investigator described how they “*had one case in particular that took 11 months, and I think every single week [the family was] emailing [them] questions.”* One death investigator described the challenge of such long wait times in cases where SCDs are attributable to heritable cardiac conditions. They said *“if I know that this family carries Brugada Syndrome, and I’m not going to get back to them for a year and a half, but I know the results, I think ethically there’s a bridge there. I [can’t] tell them and know it for months, and have another child drop dead or another family member drop dead, ethically that’s a huge void. Like a huge error on our part, to not tell them as soon as we have that information.”*

Another challenge brought forth by a death investigator in Ontario, was the necessity for families needing to request in writing their wish to receive copies of the final death investigation reports from each regional office within the Office of the Chief Coroner of Ontario. The participant went on to explain that the reports cannot be released by the death investigators themselves, and considered this a critical barrier for families who may not be aware that requesting the final report is their responsibility.

### Theme 3. How can communication improve

Death investigators offered numerous recommendations for future initiatives that may improve their communication with families, while simultaneously improve the efficiency of the system. A participant suggested sending a written letter to families of SCD victims to help them to both remember information and relay the potential risk to other family members: “*Because I mean, when they’re getting this information, it may be all a buzz to them, they could be in the grocery store when I call. They could be you know anywhere or not really understand. And then have to pass the information onto their partner or their children and they don’t really know what to tell them. You know, a standard letter that could go out and explain to them what it means in plain English.*” Others spoke about the importance of involving the family physician in the communication process as they can explain the information in the death investigation reports and initiate any referrals: “*Sometimes the family doctor will initiate it [the referral] and sometimes the family does and sometimes it’s just because they want to sit down with the family doctor and have them read and explain it to them*.”

To improve the timeliness of communicating the potential risk for heritable cardiac conditions, one suggestion was the automation of current systems, such as ensuring cases get “*flagged for follow up with genetics”* or adding a “*centralized reminder system”* to ensure cases are not delayed or, at worst, forgotten. Another death investigator suggested their institution should create new processes or even fund new role to support their investigations “*And so, I think that if the office feels that it’s really important for us to be doing genetic follow up with cases like this, there should be someone like [the family genetic care associate]], who is more actively engaging with us, right from the get-go and helping us with the investigation, so that we do things properly and we don’t miss things.”*

### Data Integration

The quantitative survey results allowed us to appreciate the various communication strategies that are used (phone, in person, text, etc), whether alone or in combination. Through the interviews, we gained a deeper appreciation as to why these various strategies are used and for what purposes (to inform the family about the cause of death; to protect the living by adequately informing family members about their risk for heritable cardiac conditions; and to recommend the referral of family members for cardiac follow-up and screening). The survey also revealed that there are challenges with suspected heritable cardiac cases, more so than other cases (higher frequency of communication, lengthier investigations, etc). The qualitative data revealed the numerous barriers at play as to why these cases are more difficult (characteristics of the family; involvement of other professionals; characteristics of the investigation, access to resources, and system-level barriers). Further training or guidelines may improve the readiness of death investigators to engage in these challenging investigations and discussions and allow for improved communication between death investigators and family members of SCD victims.

## Discussion

Investigations into young SCD cases of suspected heritable cardiac conditions are often complex and lengthy, leading to unique communication dynamics between death investigators and family members of SCD victims. The strategies death investigators use to achieve their communication goals with family members may be supported or hindered by numerous factors. Characteristics of the family, involvement of other professionals, characteristics of the death investigation, death investigators’ access to resources, and system-level barriers may all shape communication. Many of the factors influencing the communication are outside of the control of the death investigator. Death investigators are often required to adapt to the needs of the family, including family dynamics, grief, and their ability to retain information. Moreover, the uncertainty involved with SCD cases, that often last months, may result in less clear communication, compared to other cases.

Although death investigators described many strategies that they used to achieve their communication goals, there are still no formal guidelines or training for them to refer to in practice.^3^ They seem to draw on their training and experiences as physicians to develop a way to communicate with families that works for them. Our team’s prior work in Ontario has found that families of SCD victims struggle to comprehend crucial information during the stressful grieving period following the death of a family member.^5^ This indicates that the current communication strategies employed by death investigators may be insufficient in adequately informing family members of the cause of death and of their own potential risk for heritable cardiac conditions. Additionally, death investigators often independently seek outside resources, extra training, or other experts to inform their communication practices with families. All-encompassing, standardized training or guidelines may improve the preparedness of death investigators to communicate with families and to achieve their communication goals.

Similar to others,^16–18^ our team’s prior research that interviewed family members of SCD victims supports many of the perspectives provided by the death investigators.^5^ We found that both family members and death investigators understand the immense role that grief and shock take up following an unexpected death, which may require adjustments to communication strategies. Additionally, family members and death investigators both reported that SCD investigations can be more lengthy and complex, particularly due to the potential for an unknown cause of death.

### Implications for clinical practice

Death investigators provided a wealth of suggestions on how the communication processes between them and SCD families could be made more effective. Some suggested that multiple modes of communication, including a written letter in addition to verbal exchanges, should be used to help families both understand and be able to refer to the cause of death and potential risk to other family members. These are similar findings to our previous work interviewing SCD families about their experiences.^5^ In that study, family members expressed that they valued a variety of communication formats, to ensure they could review details later and make sense of important information on their own terms. Other suggestions from death investigators included the importance of involving the family physician to help explain the reports and to initiate any referrals. This is an important observation, as little is known about the role of the family physician in these situations and how they contribute to the communication of SCD risk to families and facilitate cascade screening (genetic testing of family members of an individual found to have a SCD due to a genetic cause). Future research should explore the communication processes between family physicians, SCD families and death investigators, to help streamline and facilitate the referral processes.

### Strengths and Limitations

There are strengths and limitations of this study that must be considered. A strength of our study was the integration of quantitative and qualitative data at numerous levels. We were able to use the quantitative survey results to inform the interview guide, acquiring complementary and deeper explanations into the survey responses. This allowed us to probe some of the survey results and permitted a more targeted interview process, offering deeper insights into the results of the web survey. A limitation of our study was the low number of participants from Nova Scotia, in comparison to Ontario. This is to be expected, because Nova Scotia has a much smaller population than Ontario, and hence a much smaller death investigation system to meet their needs. Our overall sample represents a very large province and death investigation system, so our results may not be applicable to smaller regions and systems. Another limitation of this study was the lack of ethnic diversity. The results from this study may not accurately reflect the more ethnically diverse Canadian population. It is unknown whether the ethnic distribution of participants in this study is representative of the ethnic distribution in these death investigation systems or others. Finally, the perspectives represented in this study only represent death investigators who volunteered to share their experiences. Death investigators who declined to participate could have distinct communication goals and strategies that are not captured. Regardless, the perspectives captured provide insight into the network of strategies that death investigators use when communicating with family members of SCD victims.

### Conclusion

Death investigators use a variety of strategies to achieve their communication goals when informing family members of SCD victims about the cause of death and their own risk for heritable cardiac conditions. The strategies used may be influenced by the (1) characteristics of the family; (2) involvement of other professionals; (3) characteristics of the investigation, (4) access to resources, and (5) system-level barriers. Further training or guidelines may allow for improved communication between death investigators and family members of SCD victims, by improving the preparedness of death investigators to engage in challenging investigations and discussions.

## Data Availability

Data may be made available upon reasonable request.

## Acknowledgments

The authors wish to thank the Office of the Chief Coroner of Ontario and the Nova Scotia Medical Examiner Service for their collaboration and assistance with this project.

